# SARS-CoV-2 cell entry gene ACE2 expression in immune cells that infiltrate the placenta in infection-associated preterm birth

**DOI:** 10.1101/2020.09.27.20201590

**Authors:** Phetcharawan Lye, Caroline E. Dunk, Jianhong Zhang, Yanxing Wei, Jittanan Nakpu, Hirotaka Hamada, Guinever E Imperio, Enrrico Bloise, Stephen G Matthews, Stephen J Lye

## Abstract

COVID-19 infection during pregnancy is associated with an increased incidence of preterm birth but neonatal infection is rare. We assessed pathways by which SARS-CoV-2 could access the placenta and contribute to fetal transmission. Placentas from pregnancies complicated with chorioamnionitis (ChA), exhibited increased expression of *ACE2* mRNA. Treatment of 2^nd^ trimester placental explants with LPS, induced an acute increase in cytokine expression followed by *ACE2* mRNA. Placental ACE2 protein localized to syncytiotrophoblast, in fetal blood vessels and M1/M2 macrophage and neutrophils within the villous stroma. Increased numbers of M1 macrophage and neutrophils were present in the placenta of ChA pregnancies. Maternal peripheral immune cells (mainly granulocytes and monocytes) express the *ACE2* mRNA and protein. These data suggest that in COVID19 positive pregnancies complicated by ChA, ACE2 positive immune cells have the potential to traffic SARS-CoV-2 virus to the placenta and increase the risk of vertical transmission to the placenta/fetus.

## Introduction

A novel coronavirus SARS-CoV-2 (2019-nCoV) appeared in Wuhan, Hubei Province, China on December 30, 2019, and spread rapidly around the world (Schwartz, 2020). The acute respiratory syndrome caused by SARS-CoV-2 was named Corona Virus Disease, COVID-19 by the World Health Organization (WHO). At present there are only a few reports identifying mother to fetus/neonate vertical transmission of COVID-19 infection (Dong *et al*., 2020). Facchetti et al (2020) recently reported the presence of a placenta intervillous inflammatory infiltrate consisting of neutrophils and monocyte-macrophages expressing activation markers in patients with COVID-19 infection, and Chen et al (2020) suggested the potential for transmission of SARS-CoV-2 from an infected mother to her fetus/neonate before birth. There is limited data on whether SARS-CoV-2 infections increase the risk of complications in pregnancy (Qiao, 2020). A recent systematic review meta-analysis of 13 publications related to COVID infection in pregnancy from China suggests a high rate of maternal and neonatal complications in infected individuals with a preterm birth rate of 20% and a lower neonatal infection rate of 6% (Capobianco *et al*., 2020).

The immune system is a core component of the maternal-fetal interface. Disruption of the immune system of a pregnant woman, as in the case of bacterial infection, places the mother and fetus/neonate at risk of other infections, such as viruses, which can impact the course of pregnancy for both mother (e.g. preterm birth) (Silasi *et al*., 2015) and fetus/neonate (e.g. systemic inflammatory response syndrome) (Francis *et al*., 2019). Additionally, it has been suggested that viral infection of the placenta modifies the immune response to bacterial products by destroying the normal ‘tolerance’ to LPS and aggravates the inflammatory response, which in turn leads to preterm labor (Cardenas *et al*., 2011; Kwon *et al*., 2014). Immune cells at the maternal-fetal interface play a critical role in a successful pregnancy. In the first trimester they contribute to remodelling of the uteroplacental circulation (Hazan 2010; Smith 2009), and at term an influx of maternal peripheral monocytes into the decidua and myometrium leads to their differentiation into macrophage which generate inflammatory mediators that contribute to the initiation of labour (Hamilton *et al*., 2012; Shynlova *et al*., 2020). However, these cells also contribute to pathologies of pregnancy. For example, in cases of acute chorioamnionitis bacterial infection of the fetal membranes and/or placenta results in an increased influx of maternal neutrophils and monocytes into these tissues where the latter differentiate into classical activated inflammatory macrophage (Kim *et al*., 2015; Cappelletti *et al*., 2020). The potential adverse consequences to the fetus of placental infection and sensitization to maternal-fetal viral transmission and the mechanisms by which this might occur require further investigation.

SARS-CoV-2 cell entry is mediated through angiotensin-converting enzyme 2 (*ACE2*), a protease responsible for the conversion of Angiotensin 1-8 (Ang II) to Angiotensin 1-7 (Ang 1-7) (Vickers *et al*., 2002; Jia *et al*., 2005). Interactions were identified between the SARS-CoV spike protein and its host receptor ACE2, which regulate both the cross-species and human-to-human transmission of SARS-CoV (Wan *et al*., 2020), and SARS-CoV-2, and share the same receptor, ACE2 (Zhang *et al*., 2020a). The SARS-CoV spike protein is known to bind ACE2 on the surface of the host cell via its subunit S1, and then fuse to viral and host membranes through subunit S2. Viral attachment is facilitated by two domains in subunit S1, which are from different coronaviruses (Li, 2016). It is currently unknown if SARS-CoV-2 binds in this same fashion. The expression and distribution of ACE2 has been reported in human fetal heart, kidneys, liver, lung, and placenta (Crackower *et al*., 2002; Hamming *et al*., 2004; Pringle *et al*., 2011a). *ACE2* mRNA expression has also been found in early gestation placenta (Pringle *et al*., 2011a; Bloise *et al*., 2020). In addition, ACE2 protein levels were found to be abundant in the syncytiotrophoblast layer (STB) and villous stroma, while less intense in the trophoblast layer (CTB) (Pringle *et al*., 2011a). The STB provides the interface between the maternal blood, containing immune cells, and the extraembryonic tissues and fetus (Knöfler *et al*., 2019; Facchetti *et al*., 2020). Interestingly activated macrophages in patients with heart failure have been shown to express high levels of the SARS-CoV-2 receptor, *ACE2* (Keidar *et al*., 2005). Furthermore, in patients with chronic obstructive pulmonary disease (COPD) activation of neutrophils, NK cells, Th17 cells, Th2 cells, Th1 cells, dendritic cells, and TNFα secreting cells can be induced by overexpression of ACE2 leading to a severe inflammatory response (Li *et al*., 2020a). Since these immune cells can potentially engage the SARS-CoV-2 virus, they may also represent a reservoir for the virus and a vector for the virus to infect the placenta/fetal membranes in cases of chorioamnionitis.

We hypothesize that the presence of an intrauterine bacterial infection will activate peripheral maternal macrophage and neutrophils and cause them to target the uterus and intrauterine tissues. If these immune cells express the SARS-CoV-2 cell entry protein, ACE2, then these cells have the potential to transport virus to the placenta and thereby increase the risk of placental infection and vertical transmission of the virus to the fetus.

## Results

### Chorioamnionitis and LPS exposure are associated with increased placental *ACE2* expression and an inflammatory cytokine/chemokine response

Placental *ACE2* mRNA expression was significantly increased (*P*<0.01) in pregnancies complicated with PTB in the presence of ChA compared with PTB alone or with uncomplicated spontaneous vaginal delivery (SVD) or not in labour (elective caesarean section, ELCS) term pregnancies (Figure 1A). Placental mRNA expression of *CCL2* was also significantly increased in ChA (*P*<0.05) compared to all other groups but there was no significant increase in *IL6/8* mRNA expression (Figure 1C-D).

**Figure 1.**
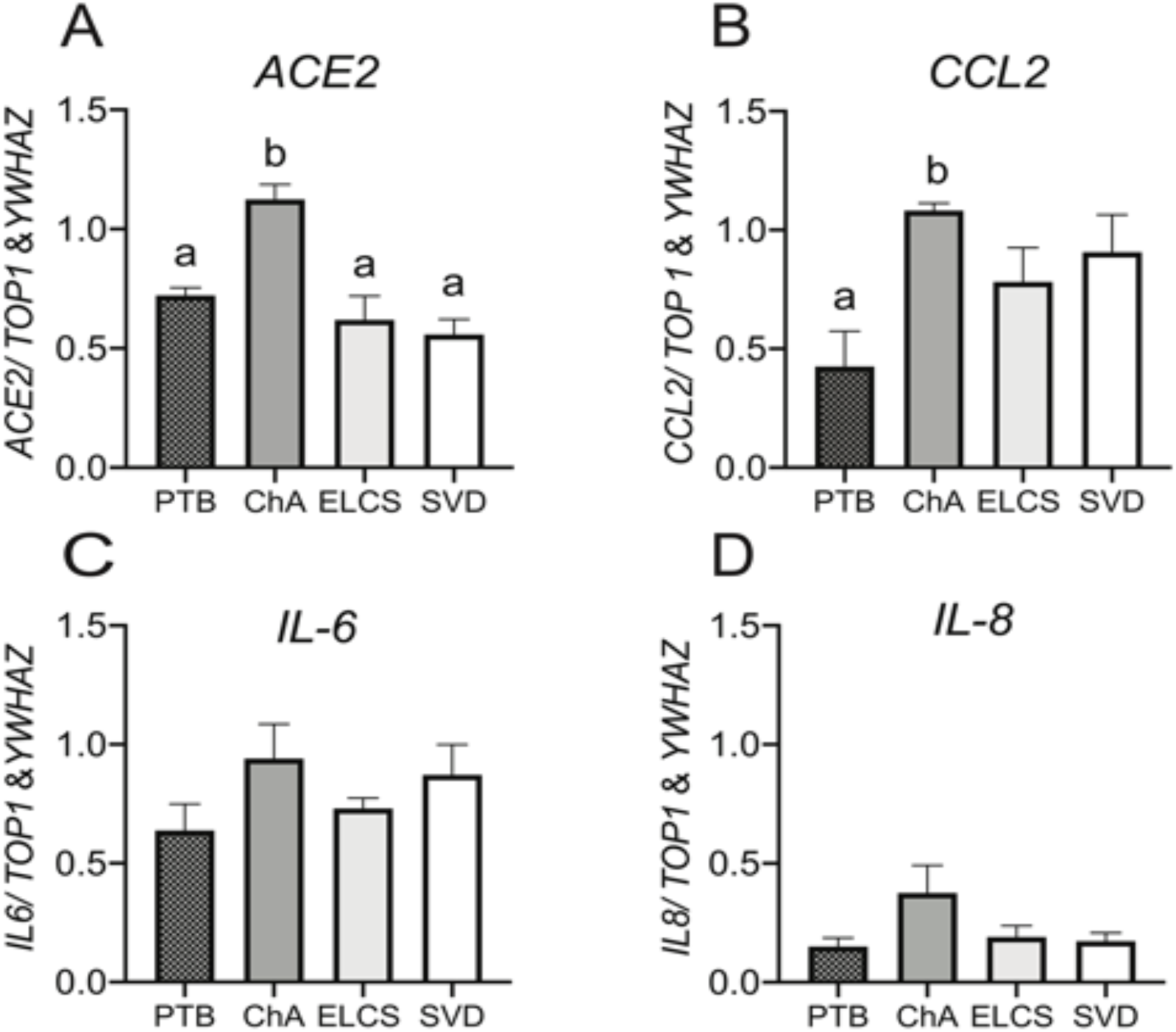
*ACE*, and chemokine mRNA expression is increased in placenta from preterm labor with chorioamnionitis (ChA). Bar charts show the relative levels of *ACE2* **(A)**, *CCL2* **(B)**, *IL-6* **(C)**, and *IL-8* **(D)** mRNA in placentas from PTB, ChA, SVD and ELCS deliveries, assessed by real time qPCR. Data are normalized by the geometric mean of *TOP1* and *YWHAZ* (reference genes), N=6-7/group. Statistical differences were tested by Tukey’s multiple comparisons test. Data are presented as mean ± SEM. Different letters indicate a difference between groups of p<0.05.

Treatment of 2^nd^ trimester placental explants with LPS (10 μg/mL) from elective terminations of healthy normal pregnancies resulted in a rapid (4 hour) significant increase in placental expression of cytokine (*IL-6/8* and *TNFα* mRNA; *P*<0.05) (Figure 2D-E), as well as higher (but non-significant expression of the chemokine *CCL2* (P=0.06)) (Figure 2B). Cytokine/chemokine expression returned to basal levels 24 hours post-LPS treatment, at which time expression of *ACE2* mRNA in the placental explants increased significantly (p<0.05) (Figure 2A).

**Figure 2.**
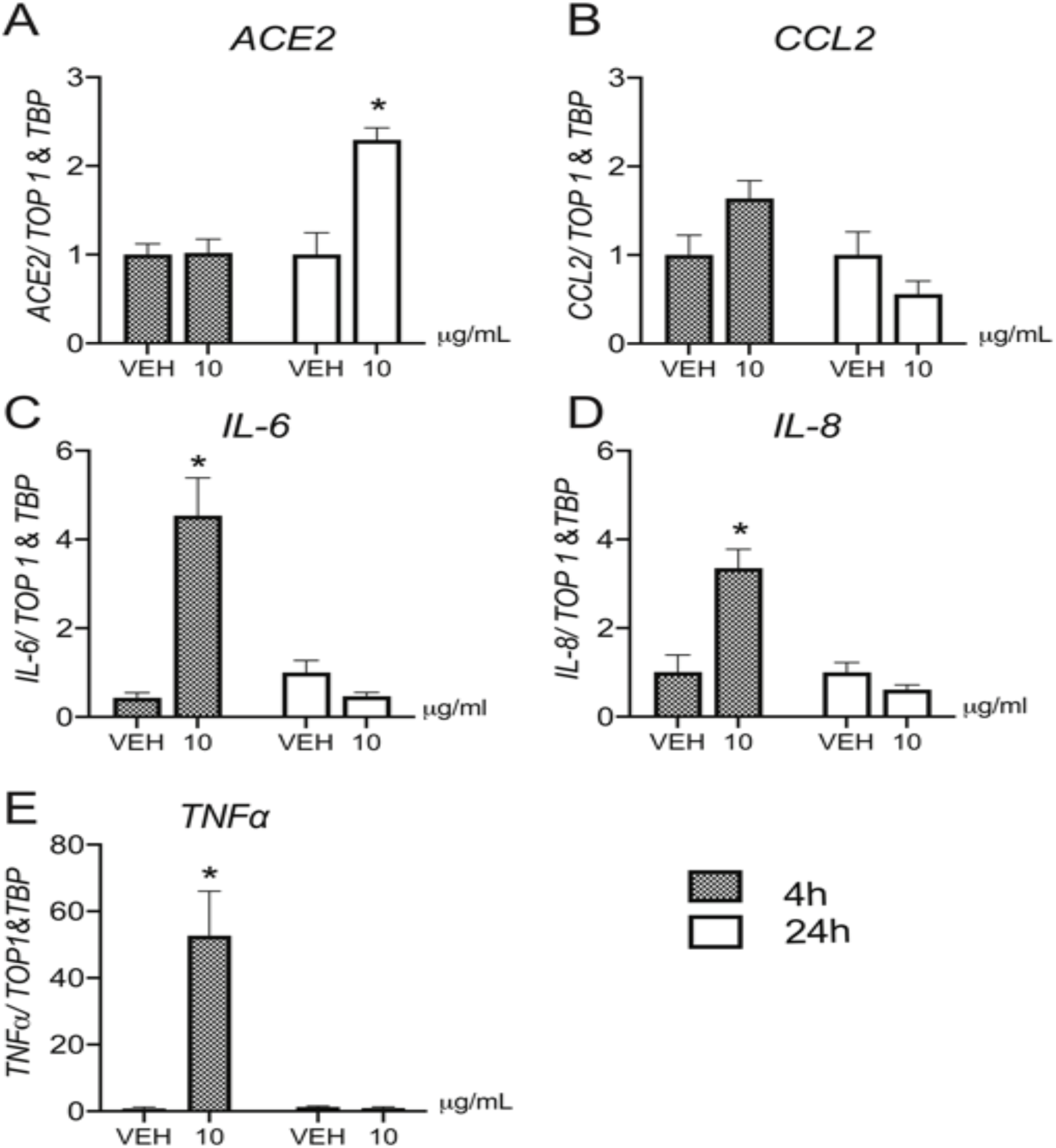
Effect of LPS on *ACE2, CCL2, IL-6/8* and *TNFα* RNA expression in second trimester human placental explants. Second trimester placental explants were treated with LPS (10 µg/mL), or vehicle for 4 and 24 hours and mRNA levels of *ACE2* **(A)**, *CCL2* **(B)**, *IL-6* **(C)**, *IL-8* **(D)** and *TNFα***(E)**, were quantified by qPCR (n=5/group). Data are normalized by the geometric mean of *TOP1* and *TBP* (reference genes). Data are expressed as means ± SEM. Statistical differences were tested using a paired *t*-test. *p<0.05, versus vehicle.

In contrast to the elevated expression of *ACE2* mRNA, we did not detect any increase in ACE2 protein within the placentas of preterm pregnancies associated with ChA, although the levels of ACE2 protein in preterm placentas (with or without ChA) were significantly higher (p<0.05) than in placentas at term in the presence or absence of labour (Figure 3B).

**Figure 3.**
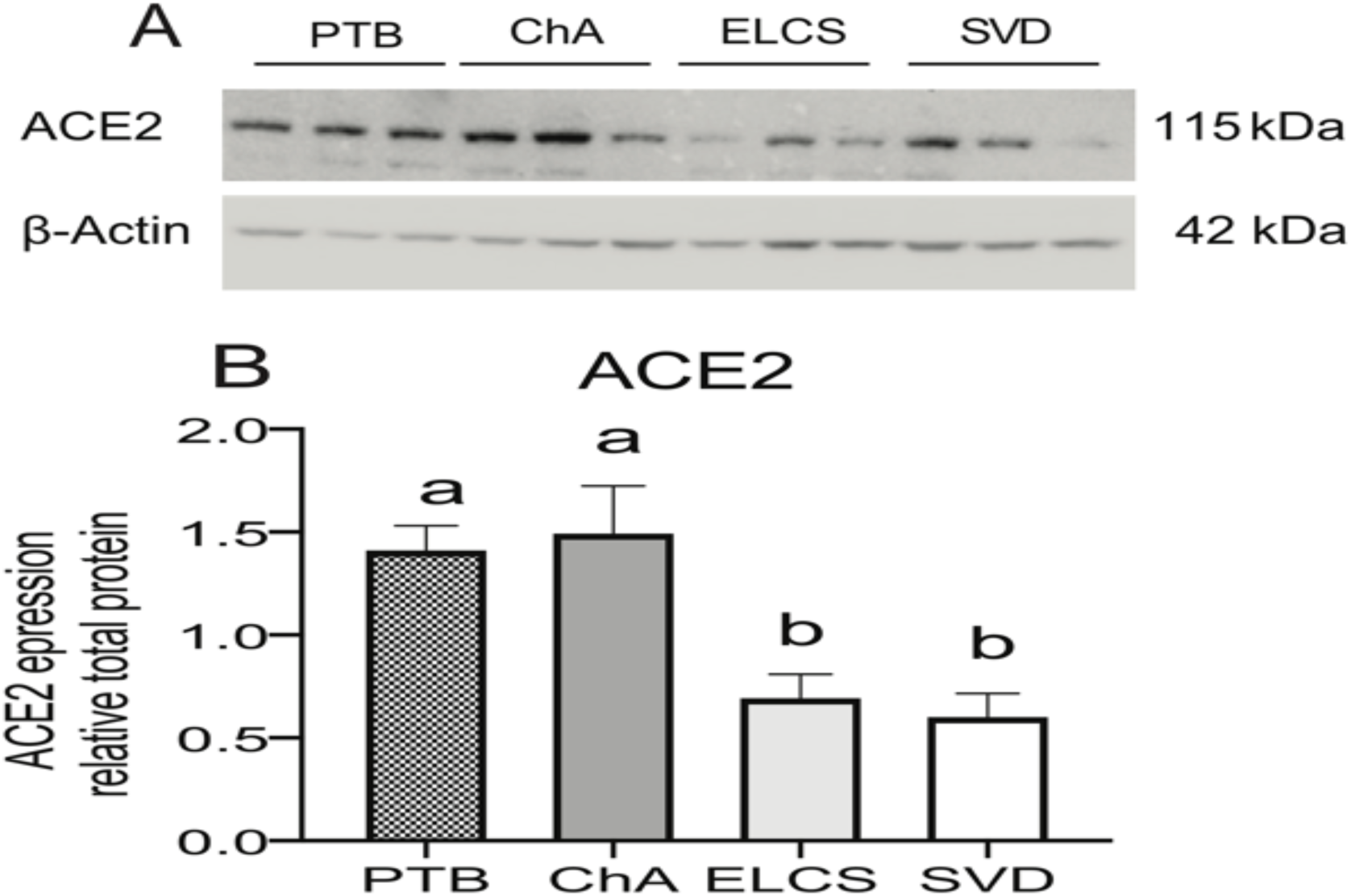
Placental ACE2 protein levels are increased in PTB with/without chorioamnionitis (ChA) as compared to term. Protein lysates of placentas from PTB, ChA, ELCS and SVD deliveries were assessed for level of ACE2 protein by western blotting. **(A)** Representative western blot images and **(B)** densitometric analysis of ACE2 protein level, normalized by *β*-actin (loading control for protein), N = 6/group. Statistical differences were tested by Tukey’s multiple comparisons test. Data are presented as mean ± SEM. Different letters if p<0.05.

Since ACE2 total protein was not increased in ChA placentas, we next determined the localization of ACE2 protein in the placentas from complicated and uncomplicated pregnancies. This analysis revealed expression of ACE2 in the syncytiotrophoblast (STB), which is the fetal tissue in direct connection with maternal blood and in the fetal blood vessels within the placental villous core. We also observed the presence of ACE2 immunoreactivity in histologically-identified macrophages within the placental villous stroma and in association with perivascular cells surrounding the fetal blood vessels and in some fetal blood cells (Figure 4A). ACE2 expression was also present in extravillous trophoblast (EVT) (Figure 4B) and in decidual stromal cells within the placental basal plate (Figure 4C), as well as immune cells within maternal blood present in the intervillous space (Figure 4D).

**Figure 4.**
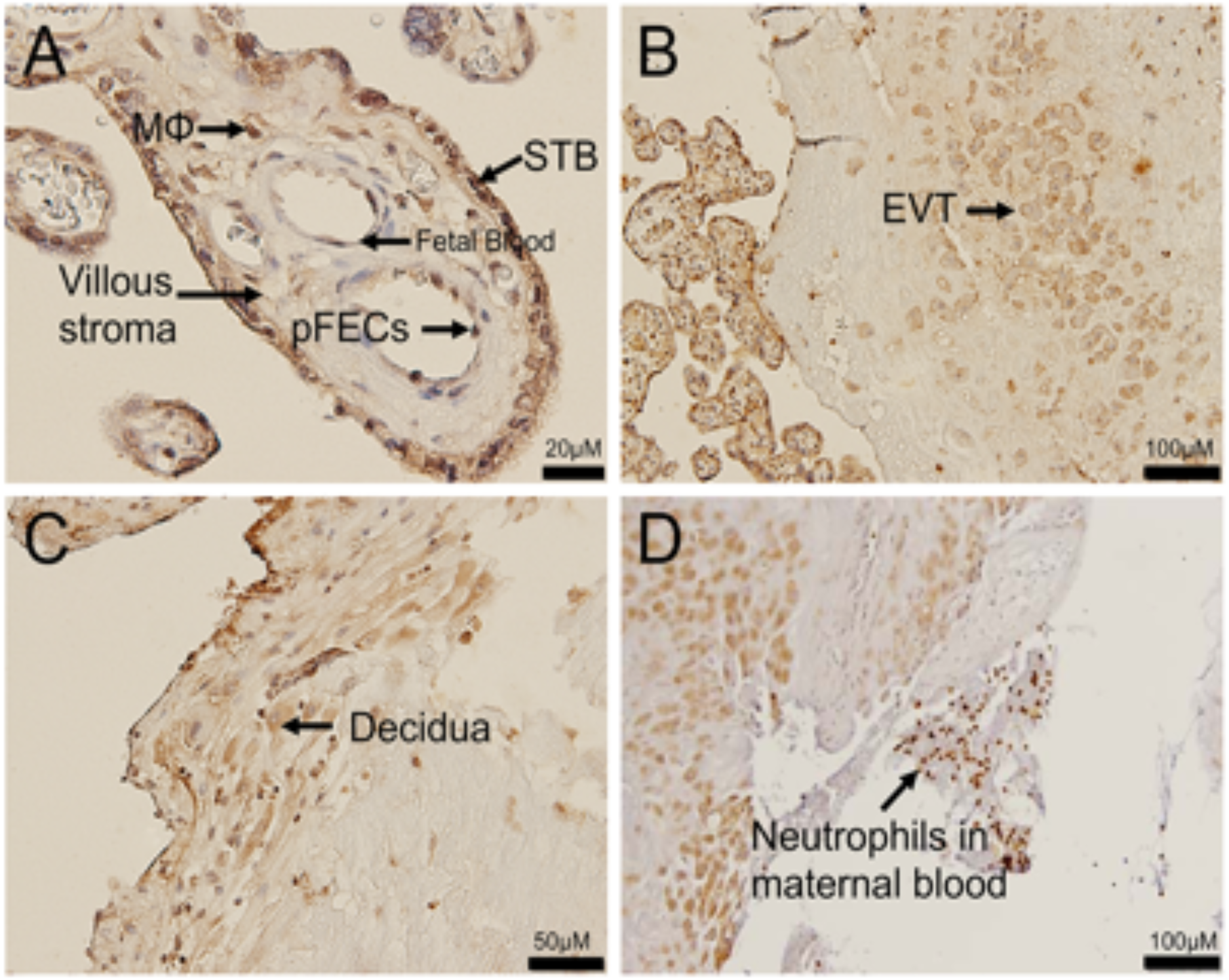
ACE2 localization in the placenta and in maternal intervillous immune cells. Representative images of ACE2 immunostaining in a placenta from an uncomplicated PTB delivery (**A)** ACE2 staining was localized predominantly in the syncytiotrophoblast layer (STB), fetal blood vessel (pFECs) and fetal hoefbauer cells (MF) of the placental villi. (**B)** ACE2 also localised to extravillous trophoblast (EVT) within the basal plate of the placenta and in **(C)** decidual stromal cells. **(D)** ACE2 expression in maternal neutrophils of maternal blood clot adjacent to the decidua, Sections were counter-stained with hematoxylin. N=6/group. Scale bar represents 100μm.

The finding of ACE2 expression in histologically-identified immune cells within the placenta led us to undertake further characterization of these cells in placentas from pregnancies complicated by PTB, in the presence or absence of ChA, as well as term pregnancies with and without the onset of labour. We used monoclonal antibodies specific for CD68 (which detect the macrophage population); MHC-II - a marker of M1 macrophage; CD206 - a marker of M2 macrophage; and neutrophil elastase (NE) - a marker of neutrophils. Representative images of immunohistochemical staining revealed expression of ACE2 was localized to syncytiotrophoblast of all pregnancy groups (Figure 5 A-D) with a reduced intensity of staining in term groups than in preterm groups. ACE2 staining was also apparent in stromal cells which resembled macrophage and neutrophils (shown in Figure 5 E-T). Staining with CD68 revealed the presence of macrophage within the placental villous of all pregnancy groups (Figure 5 E-H) and further analysis identified these to be both M1 (inflammatory) and M2 (angiogenic) sub-types. (Figure 5 I-P). The M1 macrophage were present throughout the stroma as well as being associated with fetal blood vessels. Staining with NE detected the presence of neutrophils within the placental vasculature and villi of all pregnancy groups (Figure 5 Q-T). The distribution of these immune cells did not appear to be equal across the patient groups, therefore we sought to quantify their number using Image analysis. Total macrophage numbers were similar in ChA and PTB control groups however, both of these groups had higher numbers of macrophage than the term groups (SVD, ELCS; (p<0.01) (Figure 5U). In contrast, the number of M1 macrophages was significantly increased (p<0.001) in ChA patients compared to PTB, SVD and ELCS groups (Figure 5V). Whereas, the number of M2 was significantly higher (p<0.001) in PTB patients compared to ChA, SVD and ELCS groups (Figure 5W). Quantification of neutrophil numbers across patient groups revealed significantly higher numbers in ChA pregnancies than in the PTB, SVD or ELCS groups (Figure 5X; p<0.001).

**Figure 5.**
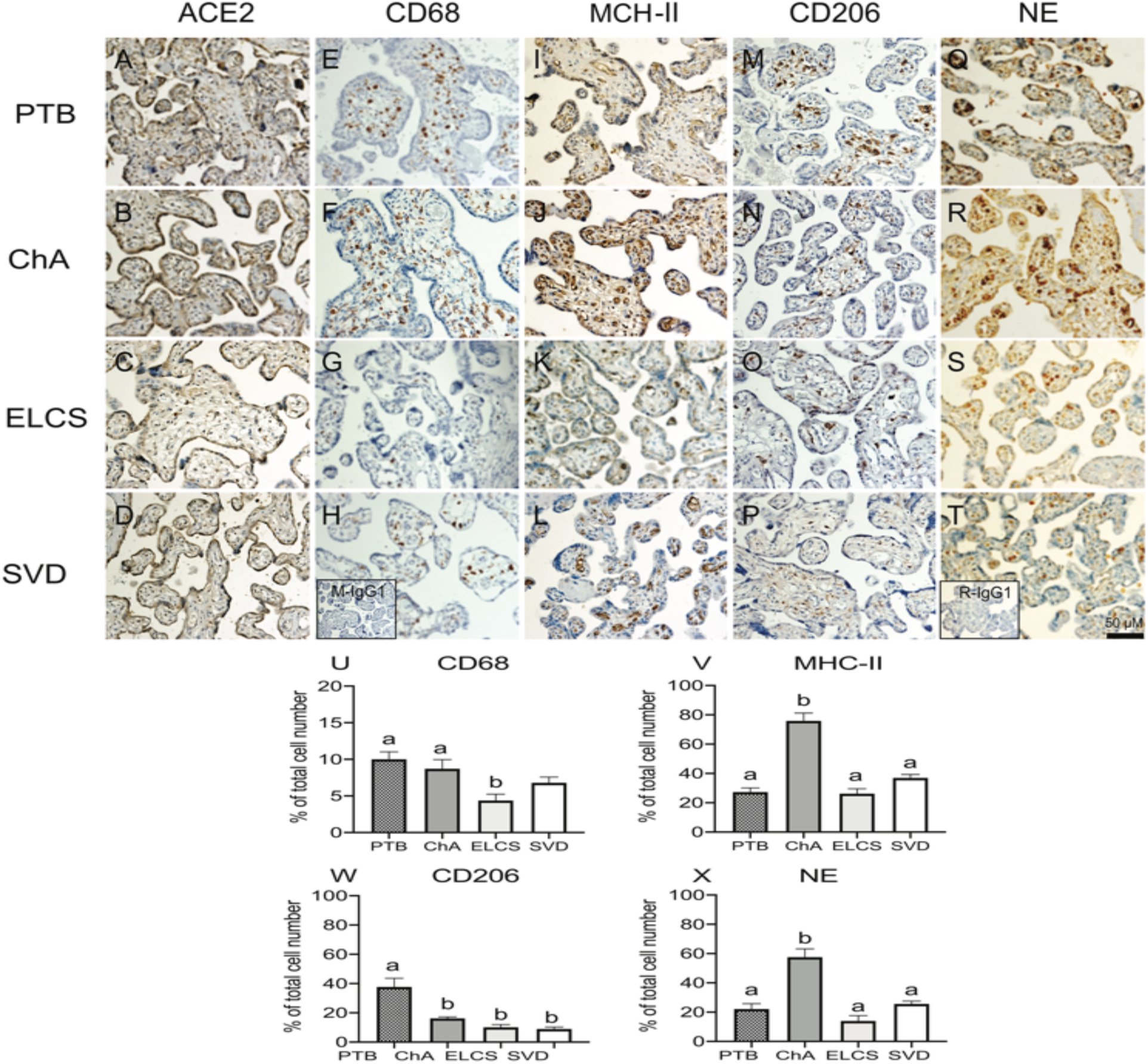
Localization and quantitation of placental ACE2, CD68 (macrophage marker), MHC-II (M1 marker), CD206 (M2 marker) and neutrophil (NE) staining in complicated and uncomplicated pregnancies. Representative images of ACE2 **(A-D)**, M1 **(MHCII E-H)**, M2 **(CD206 I-L)**, CD68 **(M-P)**, and NE **(Q-T)** staining in preterm and term placentas (N=6 in each group). Inserts (bottom left) in H mouse IgG1 and T rabbit IgG1 isotype control staining. Scale bar represents 50μm. Image analysis quantitation of positively stained immune cells as a proportion of the total cells number was performed on a randomly selected 5% of the total tissue area of each placental section. (**U)** CD68 (**V)** M1 **(W)** M2 **(X)** NE. Statistical differences were tested by Tukey’s multiple comparisons test. Data are presented as mean ± SEM. Different letters represent values significantly p<0.05.

Next we determined whether ACE2 is localized within infiltrated immune cell populations in the ChA placenta, by undertaking co-localization immunofluorescence analysis of ACE2 and CD68 (macrophage), MHC-II (M1), CD206 (M2) and NE (neutrophil) markers. Co-localization of ACE2 in macrophages (M1 and M2) and neutrophil was detected, confirming our previous histological findings that infiltrated macrophages within the placenta express ACE2 and also identifying infiltrating placental neutrophils as expressing ACE2. Immune cells expressing ACE2 were localized in the villous stroma and adjacent to fetal blood vessels. Only a subset of each of immune cells expressed ACE2 (Figure 6 A-L).

**Figure 6.**
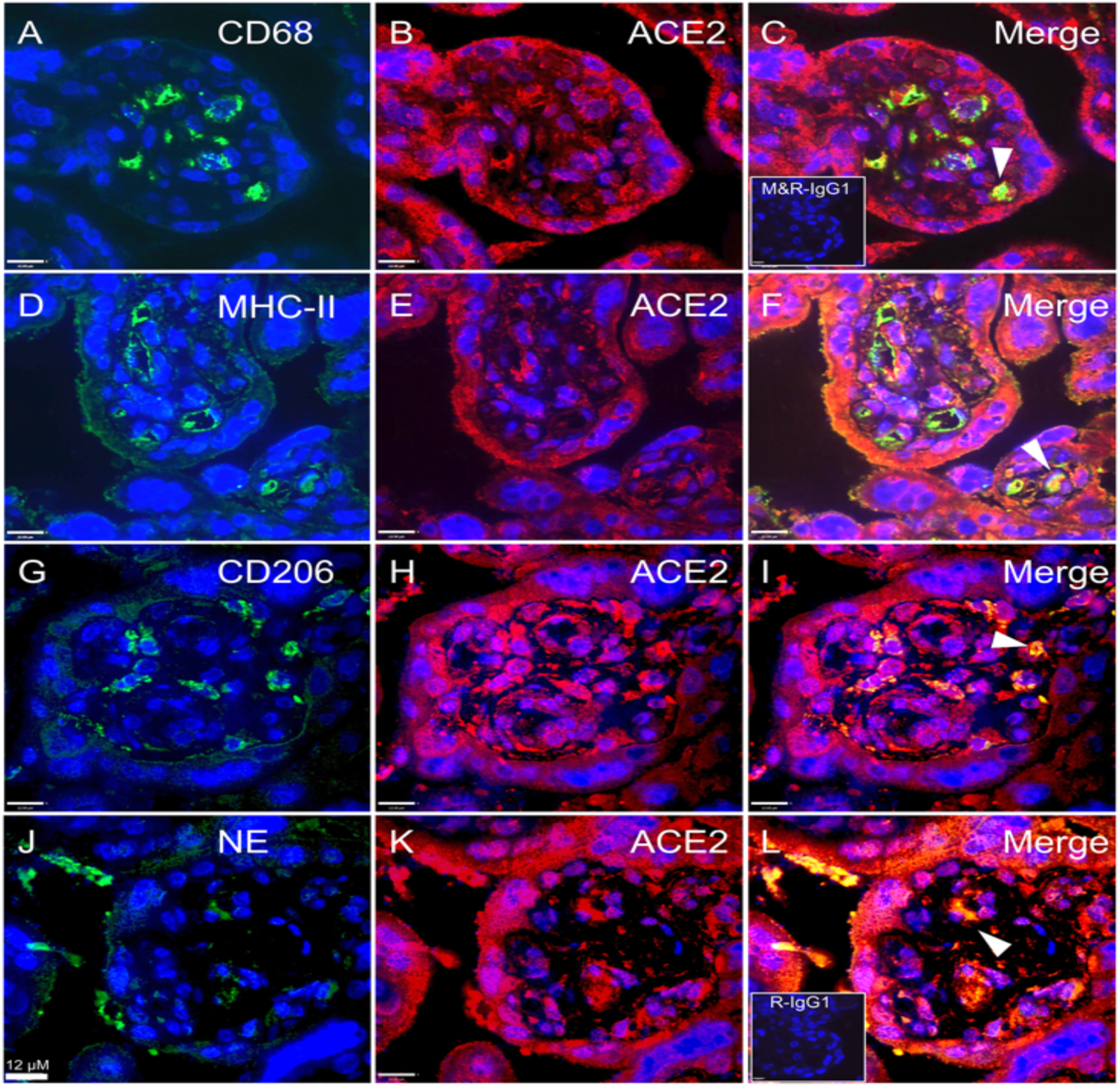
Co-localization of SARS-CoV-2 associated cell-entry protein, ACE2, within specific immune cell populations in ChA placenta. Representative images of ACE2, CD68, M1, M2 and NE staining in immunofluorescence (ACE2; red color and M1, M2, macrophage and NE; green color), macrophage co-staining with ACE2 **(A-C**) M1 co-staining with ACE2 **(D-F)**, M2 co-staining with ACE2 **(G-I)** and neutrophil co-staining with ACE2 **(J-L)**. Co-localization of ACE2 and markers of macrophage and neutrophil confirmed the ACE2 localization within placental macrophages and neutrophils. ACE2 expressing immune cells were present in the villous stroma and adjacent to fetal blood vessels. Only a subset of each of immune cells expressed ACE2. Arrows show ACE2 staining within CD68, M1, M2 and NE stained cells. Sections were counter-stained with DAPI (blue color) or co-staining (yellow color). Inserts (bottom left) in **(C)** mouse and rabbit IgG1 isotype control and **(L)** rabbit IgG1 isotype control staining. N=3/group. Scale bar represents 12 μm.

### Expression of *ACE2*/ACE2 in circulating maternal immune cells

Expression of *ACE2* mRNA was detected in circulating granulocytes, monocytes and lymphocytes from pregnant women, though levels were quite variable and were not significantly different across cell types (Figure 7A). Flow cytometry also revealed that ACE2 was expressed in fractions of circulating granulocytes, monocytes and neutrophils. A greater fraction of circulating granulocytes were found to be positive for ACE2 than lymphocytes, but not monocytes (p<0.05; Figure 7B).

**Figure 7.**
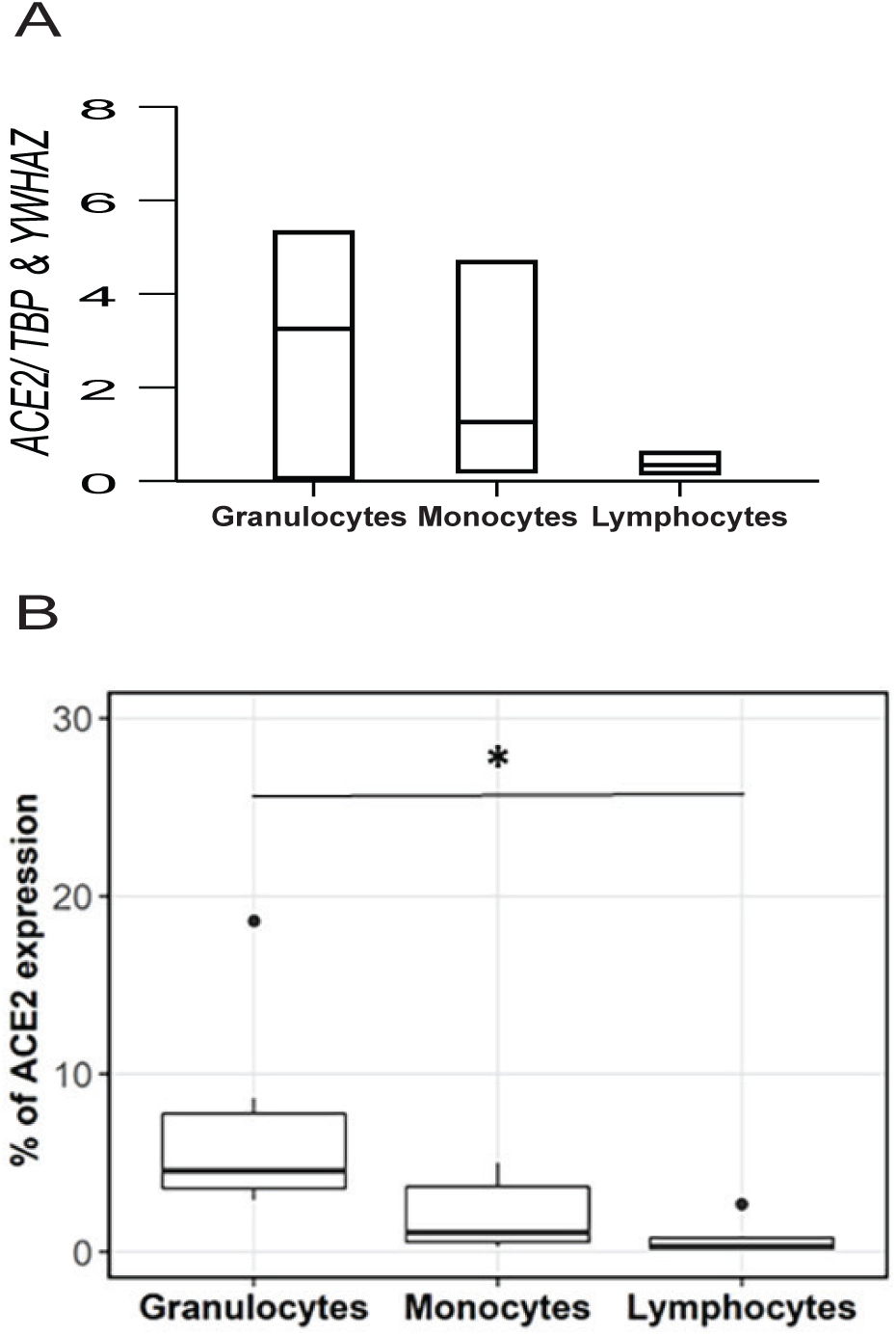
The surface expression of ACE2/*ACE2* on circulating granulocytes, monocytes and lymphocytes in pregnant women. **(A)** The relative expression of *ACE2* mRNA on granulocytes, monocytes and lymphocytes in maternal peripheral blood. There are no different between all of group (N=4/group). Statistical differences were tested by Tukey’s multiple comparisons test. Data are presented as mean ± SEM. **(B)** Peripheral human blood cells were stained with anti-human ACE2 and CD45 antibodies. The percentage of ACE2+ cells was calculated according to the gating of viable CD45+ granulocytes, monocytes and lymphocytes. Statistical analyses were performed by R software (3.4.3). Multiple comparisons were conducted by Kruskal-Wallis test and Wilcoxon test was followed to examine the statistical difference between two groups. N=6/group.*, p < 0.05.

## Discussion

In this study we provide evidence that under conditions of intrauterine infection/inflammation there is an influx of ACE2 expressing neutrophils and macrophage into the placenta. We suggest that this is a potential mechanism by which maternal-fetal vertical transmission could occur in COVID 19 infected pregnancies.

We first demonstrated that *ACE2* mRNA levels were increased in the placenta of pregnancies complicated by chorioamnionitis compared to placentas from patients of similar gestational age with no evidence of infection, or from term pregnancies (collected from non-laboring or laboring deliveries). Despite this increase in mRNA, the expression of ACE2 total protein levels (measured by WB) was not different in the placenta from patients with PTB and chorioamnionitis compared to those from PTB without chorioamnionitis, although both preterm groups expressed higher levels of ACE2 than placentas from patients at term, in the presence or absence of labour. Previously, it has been reported that there is minimal expression of ACE2 mRNA in the placenta throughout pregnancy (Pringle *et al*., 2011b; Bloise *et al*., 2020; Li *et al*., 2020b). However, we recently demonstrated that placental expression of *ACE2* mRNA is gestational-age dependent with the highest levels in early pregnancy and low to undetectable levels towards term (Bloise *et al*., 2020). Our current data suggest a similar gestational-age dependence pattern for ACE2 protein.

We found differences in the cellular distribution of ACE2 in the placenta of patients with ChA. Immunohistochemical analysis localized ACE2 to the STB layer, which is exposed to the maternal circulation as well as in the vascular endothelium of the fetal circulation. Staining was also present in extravillous trophoblast (EVT), and decidual stromal cells. Our findings are consistent with other studies that ACE2 is highly expressed in cells at the maternal-fetal interface including, STB, endothelial and perivascular cells and EVT (Valdés *et al*., 2006). Acute chorioamnionitis refers to the presence of intra-amniotic infection or “amniotic fluid infection syndrome” (Romero *et al*., 1992; Kim *et al*., 2015). Acute chorioamnionitis can also occur in the absence of demonstrable microorganisms with “sterile intra-amniotic inflammation”, and is induced by “danger signals” or DAMPs released under conditions of cellular stress, injury or death (Romero *et al*., 1992; Gomez-Lopez *et al*., 2019). The characteristic histologic and defining features of acute chorioamnionitis is a diffuse infiltration of neutrophils into the fetal membranes (Redline *et al*., 2003; Kim *et al*., 2015). All of the placentas in the ChA group were confirmed, by a placental pathologist, to be Grade 1/2, stage 2/3 with fetal inflammation of the membranes (amnion and chorion) and umbilical cord (Table 1). Importantly, we noted expression of ACE2 in immune cells within the ChA placenta. These cells were characterized to be predominantly M1 macrophage and neutrophils, many of which were found in close association or within the wall of fetal blood vessels. Quantification of the number of these cells in the placenta from all patient groups revealed, as expected, significantly greater numbers of these immune cells in pregnancies complicated by acute stage 2/3 ChA than the other pregnancy groups. A recent study has shown that both maternal and fetal neutrophils are present in the amnionitic fluid of women with a confirmed intraamniotic infection and/or inflammation (Gomez-Lopez *et al*., 2017). We did not confirm that the immune cells within the placental villi were of maternal or fetal origin in this study. However, we did also observe a maternal intervillous infiltrate of ACE2 expressing maternal neutrophils and macrophage in the ChA placenta. Further, mRNA and flow cytometric analysis of ACE2 expression by specific immune subsets in maternal peripheral blood revealed expression of the SARS CoV-2 cell entry protein in neutrophils, monocytes and lymphocytes, with the fractions of neutrophils expressing ACE2 being significantly higher than in lymphocytes, but not monocytes. The surface expression of ACE2 on these peripheral immune cells, provides a route by which the virus could enter these cells in the peripheral circulation. Since intrauterine infection (as in the case of chorioamnionitis) leads to activation and targeting of maternal immune cells (particularly neutrophils and monocytes) to the placenta (Glauser *et al*., 1991; Liu *et al*., 2018), this would provide a direct path which might facilitate the transmission of viral particles into the placental bed, and subsequently to the fetus. Thus, maternal immune cells could act as the vector for SARS CoV-2 to infect the placenta and or fetus.

**Table 1.** Clinical profile of placental tissues. BMI, (Body Mass Index); V, (Vaginal), C, (Caesarean section with no labor); CL, (Caesarean section with labor); S1, (Stage1); S2, (Stage2); S3, (Stage3); Y/N, (Yes/No).

|  | PTB<br>(N=12) | ChA<br>(N=12) | ELCS<br>(N=12) | SVD<br>(N=12) |
| --- | --- | --- | --- | --- |
| **Maternal Characteristics** |  |  |  |  |
| Maternal Age (years) | 29.6 ± 1.83<br>(21-43) | 28.5 ± 2.25<br>(17-40) | 35.8 ± 0.85<br>(34-42) | 32.4 ± 1.06<br>(27-43) |
| Labor (Y/N) | 10:2 | 11:1 | 0:12 | 12:0 |
| BMI | 22.4 ± 1.18<br>(18.26-29.75) | 23.8 ± 1.12<br>(18.30-29.75) | 22.6 ± 1.77<br>(15.24-32.44) | 23.2 ± 1.34<br>(19.08-32.91) |
| **Fetal Characteristics** |  |  |  |  |
| Gestational age (weeks) | 31.7 ± 0.93<br>(25.3-36.0) | 29.3 ± 1.01<br>(26-33) | 38.6 ± 0.30<br>(37-41) | 39.0 ± 0.32<br>(37-40.4) |
| Mode of Delivery (V:C:CL) | 2:1:9 | 5:1:6 | 0:12:0 | 12:0:0 |
| Neonatal sex (Male/ Female) | 9:3 | 8:4 | 6:6 | 7:5 |
| **Pathology Characteristics** |  |  |  |  |
| Chorioamnionitis (S1, S2, S3) | No | 1:10:1 | No | No |
| Glucocorticoids treatment | Yes | Yes | No | No |

Our data showing that LPS treatment (a model of bacterial infection and chorioamnionitis) of second trimester placental explants induces a rapid induction of chemokine/cytokines (*CCL2, IL-6/8* and *TNF*α) transcripts and *ACE2* mRNA at 24 hours provides support that placental bacterial infection might increase the opportunity for placental infection by SARS CoV-2. Since syncytiotrophoblast and placental resident immune cells express ACE2 two potential routes for viral infection might exist, one through direct interaction between virus and the outer (syncytial) surface of the placenta and one through the influx of virus containing immune cells. Moreover, since the activated immune cells infiltrating into the placenta can release proinflammatory cytokines this might further enhance placenta *ACE2* mRNA expression (Liu *et al*., 2018). Whether bacterial LPS can sensitize the placenta to infections by SARS-CoV-2 is not known, and requires further study.

There is conflicting data as to whether SARS-CoV-2 can be transmitted to the fetus through the placenta in women with COVID-19. Early reports of obstetric and neonatal outcomes of pregnant women from China with COVID-19 suggested that SARS-CoV-2 infection in pregnancy results in limited neonatal infection (Rasmussen *et al*., 2020; Zhang *et al*., 2020b). However, an increasing number of studies reporting neonatal cases with congenital or intrapartum infection. So far, 10 infants with SARS-CoV-2 infection meet these criteria based on a classification system defined by Shah et al. (Shah *et al*., 2020). Notably, a clinical case report revealed that a neonate born to a COVID-19 infected pregnant woman was tested positive for SARS-CoV-2 after 36 hours of postnatal life (Wang *et al*., 2020), which meets the criteria of intrauterine transmission (Blumberg *et al*., 2020). In addition, in one preterm pregnancy case, the baby tested negative for COVID-19 at birth, and positive 24 hours later (Zamaniyan *et al*., 2020). Three other neonates showed increased immunoglobulin M (IgM) antibodies specific to COVID19 at birth (Zeng *et al*., 2020). There is also a growing body of evidence demonstrating infection of the placental membrane and the villous trophoblasts, a recent electron microscopy study has demonstrated the presence of SARS-CoV-2 in placental STB (Algarroba *et al*., 2020) and Baud et al have reported a case of SARS-CoV-2 positivity in the placental submembranes and cotyledons together with mixed inflammatory infiltration and funicity in pregnant woman in the second trimester (Baud *et al*., 2020). In this case, placental infection of SARS-CoV-2 was confirmed by the virologic findings from specimen obtaining from the fetal surface. Previously, various pathways of viral transplacental migration were concluded based on the studies of Zika virus, including transmission from maternal blood to EVT, transcytosis via syncytiotrophoblast-expressed receptors, or spreading by infected maternal immune cells (Chen *et al*., 2020; Karimi-Zarchi *et al*., 2020).These data support our hypothesis of an increased risk of placental and fetal transmission of SARS-CoV-2 in cases of intrauterine infection/inflammation and chorioamnionitis.

In conclusion, our data show that the presence of an intrauterine bacterial infection results in the infiltration of ACE2 expressing maternal macrophage and neutrophils into and across the placental tissues (Figure 8). These ACE2 expressing immune cells have the potential to transport virus to the placenta in cases of COVID-19 infection in pregnancy and increase the risk of placental infection and vertical transmission of the virus to the fetus.

**Figure 8.**
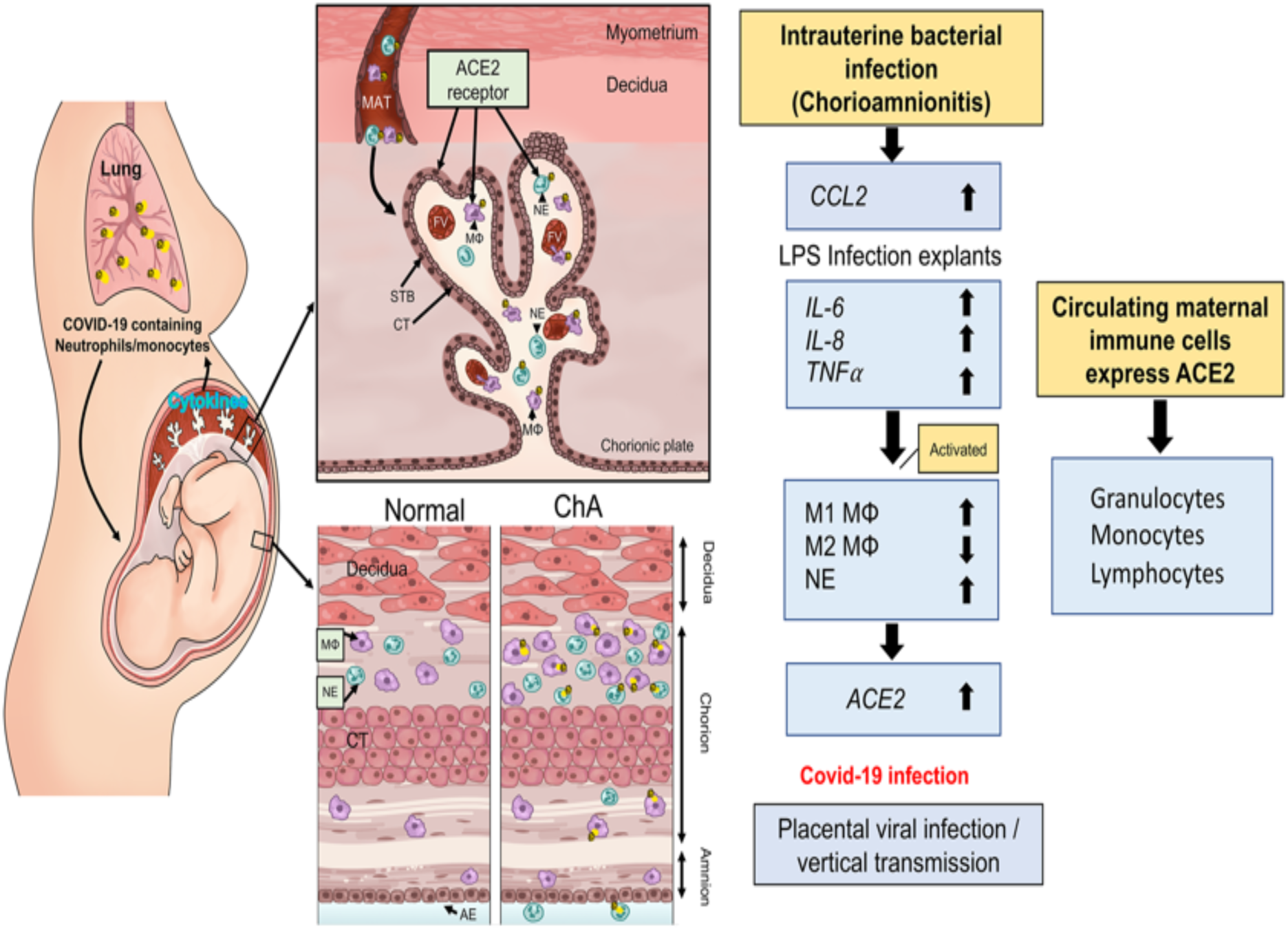
Schematic model by which maternal immune cells could bring SARS-CoV-2 to the placenta and increase risk of vertical transmission in pregnancies complicated by intrauterine infection. Intrauterine bacterial infection, such as occurs in chorioamnionitis (ChA), leads to increased expression and release of chemokine/cytokines by the placenta that in turn induce activation of maternal immune cells, notably M1 macrophage and neutrophils. These cells then target the sites of infection in the placenta/fetal membranes. Subsets of these immune cells express the SARS-CoV-2 cell entry protein, ACE2, and thus are targets for viral uptake in women infected with SARS-CoV-2. The virus may then be hypothetically transported to the placenta increasing the potential for vertical transmission of the virus to the fetus. MAT= maternal blood, FV= fetal vessel, MF= M1 macrophage, NE= neutrophil, STB= syncytiotrophoblast, layer CT= cytrophoblast.

## Methods

### Ethical Approval

This is a cross-sectional study involving placental tissue collection from pregnancies (25.3-36.0 weeks’ gestation) complicated by preterm birth (PTB; N=12) or preterm birth with chorioamnionitis (ChA; N=12) or from term pregnancies (>37 weeks’ gestation) from healthy women in labour (vaginal delivery, SVD; n=12) or not in labour (elective caesarean section, ELCS; N=12). Second trimester human placental tissues were collected at 16–20 weeks of pregnancy (N=5/group). All clinical information is depicted in Table 1.

Human peripheral blood was collected from normal pregnant women at 10-14 weeks of pregnancy (N=6). All tissues and peripheral blood were provided by the Research Centre for Women’s and Infants’ Health Bio Bank program at Sinai Health System and are collected following informed written consent (process n# 20-0101-E) and in adherence with the policies of the Sinai Health System and the University of Toronto Research Ethics Board and in accordance with the Helsinki declaration on the use of human tissues.

### Human Placental Explant Culture

Second trimester human placentae (16 to 20 weeks) from the elective termination of singleton pregnancies were used to set up the floating explant culture as described earlier (Lye *et al*., 2015). Briefly, placental specimens were placed into 1% phosphate-buffered saline (PBS) with Ca^2+^and Mg^2+^ and transported to the laboratory. Tissues were dissected into villous clusters of approximately 15 to 30 mg, and three villous explants were cultured per well in 12-well plates that contained Dulbec-co’s modified Eagle’s medium/F12, Normocin antibiotic (Invivogen, San Diego, CA), and 1 insulin, transferrin, and selenium-A (Invitrogen, Grand Island, NY) that was previously equilibrated at 8% O2 (CO2, 37 °C) for 24 hours. Explants were cultured for 24 hours and then randomly divided into treatment groups. Explants were treated with LPS from Escherichia coli (10 μg/mL L4391, Sigma-Aldrich, St. Louis, MO) or vehicle for 4 or 24 hours (Lye *et al*., 2015). Explants were then collected and stored at 80°C for total RNA.

### Immunohistochemistry

Placental tissues (N=5/6group) were processed for immunohistochemical analysis as previously described (Lye *et al*., 2019). Briefly, slides were deparaffinized, rehydrated, and subjected to heat mediated antigen retrieval with 10mM sodium citrate Ph6.0 for CD206 and neutrophil primary antibodies, or 1mM EDTA Ph9.0 for MHC-II, CD68 and ACE2. After blocking with Dako protein block (Dako, Mississauga, ON, Canada), the slides were incubated overnight at 4°C with primary antibodies: anti-rabbit ACE2 (1:200, ab15348, Abcam, Toronto, ON, Canada USA) anti-rabbit MHC-II (M1 marker) (1:400, ab180779, Abcam), anti-rabbit mannose receptor (CD206 (M2),1:200, ab64693, Abcam), anti-mouse CD68 (macrophage) (1:100, Dako), anti-rabbit neutrophil (NE)(1:100, ab21595, abcam) was added instead of the primary antibody. After incubation, the slides were washed and incubated with the corresponding biotinylated secondary antibody (1:300, 1 hour, Dako). Sections were washed in 1× PBS (3 × 5 min) and incubated with streptavidin-HRP (1 h; Dako); immunostaining was detected with the peroxidase substrate kit DAB (Dako). The slides were counterstained with hematoxylin, dehydrated in ascending concentrations of ethanol, and cover slipped with mounting medium. Visualization was undertaken with an Olympus BX61 upright, motorized microscope, and representative images were captured using an Olympus DP72 digital camera (Olympus, Tokyo, Japan. Negative controls were performed using either isotype Mouse IgG1 or rabbit IgG/IgG1 antibodies.

### Image Analysis and Quantification

Quantification of macrophages subtypes and neutrophils in placental tissues (N=5-6/group) were performed using an Olympus BX61 microscope equipped with an Olympus DP72 camera, and the newCAST software (Visopharm). Counts were performed as previously described (Dunk *et al*., 2019), using a standard protocol that assigned random counting frames covering 5% of each total masked tissue area. Brown, positively-staining cells and blue, negatively-staining cells (haematoxylin-stained) were counted at 10X magnification. A positively stained ratio was generated by dividing the total numbers of brown, positively-stained cells by the total number of cells counted in the tissue area).

### Double immunofluorescence staining

Different protocols were used for the simultaneous detection of two antigens depending on the availability of primary antibodies raised in the same or different species. For the localization of ACE2+CD68 (primary antibodies different species), immunofluorescence experiments were performed as described previously (Nadeem *et al*., 2013). In brief, ChA placental tissue slides were deparaffinized, rehydrated, and subjected to heat mediated antigen retrieval with 1mM EDTA Ph9. Autofluorescence was reduced using 0.1% Sudan Black in 70% ethanol (1 minute) and non-specific binding was blocked using 0.1% BSA, 0.3% Triton X-100 and 1% donkey serum in PBS for 1 hour. Placental slides were incubated with primary antibodies ACE2 (1:100, ab15348, Abcam), anti-mouse CD68 (1:100, Dako), anti-mouse IgG1 (Dako) and anti-rabbit Igg1 (ab171870, Abcam) added as an isotype control overnight at 4°C. Subsequently, ACE2 + CD68 slides were washed three times and incubated with fluorescent secondary antibodies, using either the anti-mouse Alexa 488 (1:1000) or the anti-rabbit Alexa 594 (1:1000) secondary antibodies (Thermo Fisher Scientific and counterstained with 1 μg/mL of DAPI, for 1 hour).

For the localization of ACE2+MHC-II (M1), ACE2+CD206 (M2) and ACE2+Neutrophil elastase (NE) (primary antibodies the same species), immunofluorescence experiments were performed as described previously (Salio *et al*., 2005; Dunk *et al*., 2018; Choudhury *et al*., 2019). In brief, ChA placental slides were deparaffinized, rehydrated, and subjected to antigen retrieval with 1mM EDTA Ph9.0. Autofluorescence was reduced using 0.1% Sudan Black in 70% ethanol (1 minutes) and non-specific binding was blocked using 0.1% BSA, 0.3% Triton X-100 and 1% donkey serum in PBS for 1 hour. Placental tissues slides were incubated with primary antibodies ACE2 (1:100, ab15348, Abcam) overnight at 4°C. Subsequently, ACE2 slides were washed three times and incubated with fluorescent secondary antibodies using anti-rabbit Alexa 594 (1:1000) (Thermo Fisher Scientific for 1 hour). Slides were washed three times and blocked with 0.1% BSA, 0.3% Triton X-100 and 5% goat serum in PBS for 1 h. Slides were incubated with primary antibodies anti rabbit-MHC II (M1) (1:100, Abcam), anti-rabbit mannose receptor (CD206, M2,1:100, Abcam) anti-rabbit NE (1:100, Abcam) and anti-rabbit IgG1 (Abcam) added as an isotype control overnight at 4°C. Slides were incubated with the corresponding biotinylated secondary antibody (1:300, 1 h, Dako) then washed three times and incubated with Streptavidin-Alexa488 (1:2000, Thermo Fisher Scientific and nuclei were stained with 1 μg/mL of DAPI, for 1 h). All slides from the primary antibodies as the same or different species were washed three times. Fluorescent microscopy was performed using a spinning disc confocal microscope at various magnifications.

### Quantitative Real Time PCR (qPCR)

Total RNA was isolated from flash frozen, stored in −80C placental specimens using the RNeasy Plus Universal Mini Kit (73404, Qiagen, Toronto, ON, Canada), as previously described (Lye *et al*., 2019). RNA concentration and purity were assessed using a NanoDrop1000 Spectrophotometer (Thermo Scientific). RNA was reverse transcribed into cDNA using the iScript Reverse Transcription Supermix (Bio-Rad). *ACE2, IL-6, IL-8, TNFα* and *CCL2* mRNA levels were measured by qPCR using SYBR Green reagent (Sigma-Aldrich) and the CFX 380 Real-Time system C1000 TM Thermal Cycler (Bio-Rad), with the following cycling conditions: initial denaturation at 95 °C (2 min) followed by 39 cycles of denaturation at 95 °C (5 s) and combined annealing and extension at 60°C (20 s). Gene expression was normalized to the geometric mean of selected reference genes, including DNA topoisomerase 1 (*TOP1*) and the zeta polypeptide (*YWHAZ*) which had stable expression across pregnancies complicated by PTB or ChA or from SVD or ELCS groups. *TOP1* and TATA-box binding protein (*TBP*) were used in studies of LPS treatments in second trimester explants. The relative expression of target genes was calculated by the 2-ΔΔCT method (Livak & Schmittgen, 2001). The primer sequences of all the assessed genes are shown in Table 2.

**Table 2.**
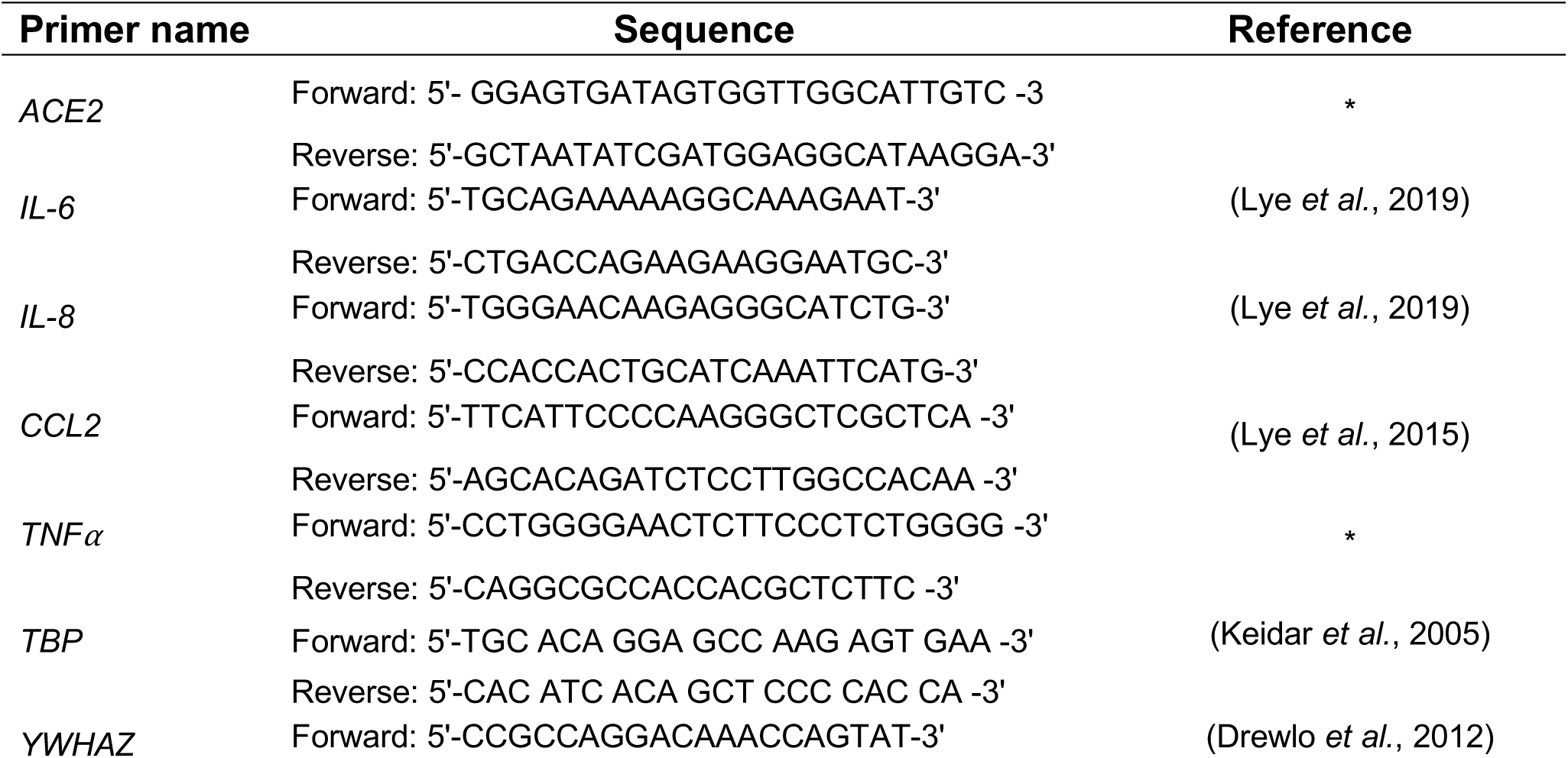
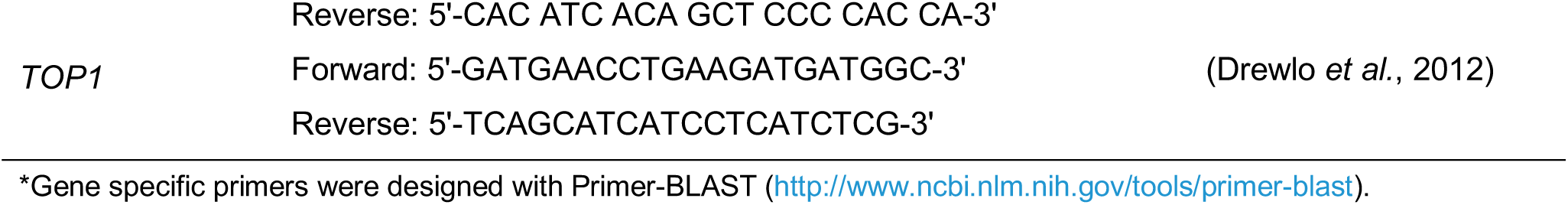
List of primers used in this study.

### Immunoblotting

Western blot analysis was conducted as previously described (Lye *et al*., 2019) Briefly, protein isolated from cultured cells was extracted by sonication using lysis buffer (1 mol/L Tris-HCL pH 6.8, 2% SDS, 10% glycerol with added protease and phosphatase inhibitor cocktail; Thermo Scientific). The protein concentration was determined with the Pierce BCA Protein Assay kit (Thermo Scientific). Proteins were separated by electrophoresis (30 μg 100 V, 1 h) using SDS polyacrylamide gels (8%). Proteins were then transferred (10 min) to polyvinylidene fluoride (PVDF) membrane using Trans-Blot Turbo (Bio-Rad). Membranes were blocked with skim milk (5%; 1 h at room temperature). The primary antibodies used were anti-rabbit ACE2 (dilution 1:1000; Abcam, ab108209, Toronto, ON, Canada) and anti-goat *β*-actin (dilution 1:2000; Santa Cruz Biotechnology, Dallas, TX, USA). Blots were incubated with primary antibodies overnight at 4 °C. The PVDF membranes were subsequently incubated for 1 h with HRP-linked anti-goat secondary antibody (GE Healthcare Bio-Science, Baie d’Urfe, QC, Canada) at concentrations of 1:10,000. Protein-antibody complexes were detected by incubating (for 5 min) the PVDF membranes with Laminate Crescendo Western HRP Substrate (Millipore, Oak Drive, CA, USA) and chemiluminescence was detected under UV by using the ChemiDoc™ MP Imaging system (Bio-Rad). The protein band intensity was quantified using Image Lab™ software.

### Immune cell isolation

Peripheral blood samples were collected prospectively into EDTA blood collection tubes from pregnant women in the first trimester (10-14 gestational weeks) during the regular antenatal visit. Monocytes and lymphocytes were isolated separately using the RosetteSep Human Total Lymphocyte Enrichment Cocktail and the RosetteSep Human Total Lymphocyte Enrichment Cocktail (both from Stemcell Technologies, BC, CA) following manufacturer’s instructions as described earlier (Paquette *et al*., 2018). RosetteSep technology uses high-density gradient centrifugation to generate a highly purified and stable leukocyte fraction RNA expression studies. Following isolation, the monocytes and lymphocytes were re-suspended in RPMI-1640 with 10% FBS. Primary human neutrophils were isolated by Histopaque double density gradient method, consisting of HISTOPAQUE^®^-1119 and HISTOPAQUE^®^-1077 (Sigma-Aldrich, MO, USA). Neutrophils were collected, washed twice in HBSS+ and spun again (Amsalem *et al*., 2014).

### Flow cytometry

Direct immunofluorescence staining was performed using peripheral blood collected from pregnant women at 10-14 weeks of pregnancy. In brief, the whole blood (50µl) was stained with LIVE/DEAD® fixable cell stain kit (L/D-violet; Invitrogen) and then incubated with human Fc block (BD Pharmingen) and incubated for 30 minutes. To detect the expression of ACE2 protein, surface staining was conducted by Alexa Fluor® 700-conjugated mouse anti-human ACE-2 (R&D Systems) and APC-H7 mouse anti-human CD45 (BD Biosciences) antibodies. Cells were washed and resuspended in stabilizing fixative buffer (BD Biosciences) to preserve immunofluorescence staining signals of human blood.

Flow cytometric data were acquired with a Gallios flow cytometer (Beckman Coulter). Data analyses were performed by FlowJo V10 (TreeStar) or Kaluza 2 (Beckman Coulter) software. Only viable CD45+ blood cells were analyzed for their ACE2 expression. Circulating lymphocytes, monocytes and granulocytes were recognized by their distinct morphological features in the forward and side scatter distribution.

### Statistical Analysis

All analyses were conducted blind to the experimental conditions. Data analyses were performed with Prism version 8 (GraphPad Software Inc., San Diego, CA, USA) qPCR data were assessed for normal distribution using D’Agostino and Pearson or the Shapiro-Wilk test; outliers were identified using “QuickCalcs” Outlier calculator program (version 7.0; GraphPad Software, Inc., San Diego, CA). Gene and protein expression, number of macrophages, and maternal immune cell were analyzed using one-way ANOVA followed by Tukey’s multiple comparisons test. Results from LPS-treated explants were assessed by paired *t*-test. Gene expression of the LPS treated explants were normalized to their respected vehicle group at both 4 and 24 hours. Differences were considered statistically significant when p< 0.05.

## Data Availability

The submission contains no supplementary material

## Author Contributions

Conceptualization: PL, CED, SGM, SJL; Funding acquisition: SJL, SGM;

Methodology: PL, CED, JZ, JN, GEI, SL; Project administration SGM, SJL; Supervision: SGM, SJL; Validation: PL, CED, JZ; Writing - original draft: PL, CED, HH, EB, JZ, YW; Writing - review & editing: CED, PL, HH, JN, YW, GEI, EB, SGM, SJL.

## Acknowledgements

The authors thank the donors, RCWIH BioBank, the Lunenfeld-Tanenbaum Research Institute, and the Mount Sinai Hospital/UHN Department of Obstetrics and Gynaecology for the human specimens used in this study (http://biobank.lunenfeld.ca). We are grateful for the assistance provided by Mrs. Anna Dorogin in optimising immunostaining, Mrs. Elzbieta Matysiak-Zablocki with the isolation of granulocytes, monocytes and lymphocytes and Dr. Oksana Shynlova with the isolation of maternal immune cell protocols.

## Additional Information

The authors report no conflict of interest.

E.B. is supported by the Coordenaçao de Aperfeiçoamento Pessoal de Nível Superior [CAPES]; finance code 001, CAPES-Print fellowship). S.J.L. is supported by a grant (FDN-143262) from the Canadian Institutes of Health Research (CIHR). S.G.M. is supported by a grant (FDN-148368) from the CIHR.

The funders had no role in the experimental design, data acquisition, analysis, and interpretation or in the manuscript writing and conception of this study. The corresponding authors had full access to all the data in the study and had final responsibility for the decision to submit for publication.

## Additional Information

Available from the authors upon reasonable request

## References

Algarroba, G.N., Rekawek, P., Vahanian, S.A., Khullar, P., Palaia, T., Peltier, M.R., Chavez, M.R. & Vintzileos, A.M. (2020) Visualization of SARS-CoV-2 virus invading the human placenta using electron microscopy. American journal of obstetrics and gynecology.

Amsalem, H., Kwan, M., Hazan, A., Zhang, J., Jones, R.L., Whittle, W., Kingdom, J.C., Croy, B.A., Lye, S.J. & Dunk, C.E. (2014) Identification of a novel neutrophil population: proangiogenic granulocytes in second-trimester human decidua. Journal of immunology (Baltimore, Md. : 1950), 193, 3070–3079.

Baud, D., Greub, G., Favre, G., Gengler, C., Jaton, K., Dubruc, E. & Pomar, L. (2020) Second-Trimester Miscarriage in a Pregnant Woman With SARS-CoV-2 Infection. Jama, 323, 2198–2200.

Bloise, E., Zhang, J., Nakpu, J., Hamada, H., Dunk, C.E., Li, S., Imperio, G.E., Nadeem, L., Kibschull, M., Lye, P., Matthews, S.G. & Lye, S.J. (2020) Expression of severe acute respiratory syndrome coronavirus 2 cell entry genes, angiotensin-converting enzyme 2 and transmembrane protease serine 2, in the placenta across gestation and at the maternal-fetal interface in pregnancies complicated by preterm birth or preeclampsia. American journal of obstetrics and gynecology. S0002-9378(20)30884-X.

Blumberg, D.A., Underwood, M.A., Hedriana, H.L. & Lakshminrusimha, S. (2020) Vertical Transmission of SARS-CoV-2: What is the Optimal Definition? American journal of perinatology, 37, 769–772.

Capobianco, G., Saderi, L., Aliberti, S., Mondoni, M., Piana, A., Dessole, F., Dessole, M., Cherchi, P.L., Dessole, S. & Sotgiu, G. (2020) COVID-19 in pregnant women: A systematic review and meta-analysis. European journal of obstetrics, gynecology, and reproductive biology, 252, 543–558.

Cappelletti, M., Presicce, P. & Kallapur, S.G. (2020) Immunobiology of Acute Chorioamnionitis. Front Immunol, 11, 649–649.

Cardenas, I., Mor, G., Aldo, P., Lang, S.M., Stabach, P., Sharp, A., Romero, R., Mazaki-Tovi, S., Gervasi, M. & Means, R.E. (2011) Placental viral infection sensitizes to endotoxin-induced pre-term labor: a double hit hypothesis. American journal of reproductive immunology (New York, N.Y. : 1989), 65, 110–117.

Chen, H., Guo, J., Wang, C., Luo, F., Yu, X., Zhang, W., Li, J., Zhao, D., Xu, D., Gong, Q., Liao, J., Yang, H., Hou, W. & Zhang, Y. (2020) Clinical characteristics and intrauterine vertical transmission potential of COVID-19 infection in nine pregnant women: a retrospective review of medical records. Lancet (London, England), 395, 809–815.

Choudhury, R.H., Dunk, C.E., Lye, S.J., Harris, L.K., Aplin, J.D. & Jones, R.L. (2019) Decidual leucocytes infiltrating human spiral arterioles are rich source of matrix metalloproteinases and degrade extracellular matrix in vitro and in situ. American journal of reproductive immunology (New York, N.Y. : 1989), 81, e13054.

Crackower, M., Sarao, R., Oudit, G., Yagil, C., Kozieradzki, I., Scanga, S., Oliveira-dos-Santos, A., Costa, J., Zhang, L., Pei, Y., Scholey, J., Ferrario, C., Manoukian, A., Chappell, M., Backx, P., Yagil, Y. & Penninger, J. (2002) Angiotensin-converting enzyme 2 is an essential regulator of heart function. Nature, 417, 822–828.

Dong, L., Tian, J., He, S., Zhu, C., Wang, J., Liu, C. & Yang, J. (2020) Possible Vertical Transmission of SARS-CoV-2 From an Infected Mother to Her Newborn. Jama, 323, 1846–1848.

Drewlo, S., Levytska, K. & Kingdom, J. (2012) Revisiting the housekeeping genes of human placental development and insufficiency syndromes. Placenta, 33, 952–954.

Dunk, C., Kwan, M., Hazan, A., Walker, S., Wright, J.K., Harris, L.K., Jones, R.L., Keating, S., Kingdom, J.C.P., Whittle, W., Maxwell, C. & Lye, S.J. (2019) Failure of Decidualization and Maternal Immune Tolerance Underlies Uterovascular Resistance in Intra Uterine Growth Restriction. Frontiers in Endocrinology, 10.

Dunk, C.E., Pappas, J.J., Lye, P., Kibschull, M., Javam, M., Bloise, E., Lye, S.J., Szyf, M. & Matthews, S.G. (2018) P-Glycoprotein (P-gp)/ABCB1 plays a functional role in extravillous trophoblast (EVT) invasion and is decreased in the pre-eclamptic placenta. Journal of cellular and molecular medicine, 22, 5378–5393.

Facchetti, F., Bugatti, M., Drera, E., Tripodo, C., Sartori, E., Cancila, V., Papaccio, M., Castellani, R., Casola, S., Boniotti, M.B., Cavadini, P. & Lavazza, A. (2020) SARS- CoV2 vertical transmission with adverse effects on the newborn revealed through integrated immunohistochemical, electron microscopy and molecular analyses of Placenta. EBioMedicine, 59, 102951.

Francis, F., Bhat, V., Mondal, N., Adhisivam, B., Jacob, S., Dorairajan, G. & Harish, B.N. (2019) Fetal inflammatory response syndrome (FIRS) and outcome of preterm neonates - a prospective analytical study. The journal of maternal-fetal & neonatal medicine : the official journal of the European Association of Perinatal Medicine, the Federation of Asia and Oceania Perinatal Societies, the International Society of Perinatal Obstet, 32, 488–492.

Glauser, M.P., Zanetti, G., Baumgartner, J.D. & Cohen, J. (1991) Septic shock: pathogenesis. Lancet (London, England), 338, 732–736.

Gomez-Lopez, N., Romero, R., Maymon, E., Kusanovic, J.P., Panaitescu, B., Miller, D., Pacora, P., Tarca, A.L., Motomura, K., Erez, O., Jung, E., Hassan, S.S. & Hsu, C.D. (2019) Clinical chorioamnionitis at term IX: in vivo evidence of intra-amniotic inflammasome activation. Journal of perinatal medicine, 47, 276–287.

Gomez-Lopez, N., Romero, R., Xu, Y., Leng, Y., Garcia-Flores, V., Miller, D., Jacques, S.M., Hassan, S.S., Faro, J., Alsamsam, A., Alhousseini, A., Gomez-Roberts, H., Panaitescu, B., Yeo, L. & Maymon, E. (2017) Are amniotic fluid neutrophils in women with intraamniotic infection and/or inflammation of fetal or maternal origin? American journal of obstetrics and gynecology, 217, 693.e691–693.e616.

Hamilton, S., Oomomian, Y., Stephen, G., Shynlova, O., Tower, C.L., Garrod, A., Lye, S.J. & Jones, R.L. (2012) Macrophages infiltrate the human and rat decidua during term and preterm labor: evidence that decidual inflammation precedes labor. Biology of reproduction, 86, 39.

Hamming, I., Timens, W., Bulthuis, M.L.C., Lely, A.T., Navis, G.J. & van Goor, H. (2004) Tissue distribution of ACE2 protein, the functional receptor for SARS coronavirus. A first step in understanding SARS pathogenesis. J Pathol, 203, 631–637.

Jia, H.P., Look, D.C., Shi, L., Hickey, M., Pewe, L., Netland, J., Farzan, M., Wohlford-Lenane, C., Perlman, S. & McCray, P.B.,Jr. (2005) ACE2 receptor expression and severe acute respiratory syndrome coronavirus infection depend on differentiation of human airway epithelia. J Virol, 79, 14614–14621.

Karimi-Zarchi, M., Neamatzadeh, H., Dastgheib, S.A., Abbasi, H., Mirjalili, S.R., Behforouz, A., Ferdosian, F. & Bahrami, R. (2020) Vertical Transmission of Coronavirus Disease 19 (COVID-19) from Infected Pregnant Mothers to Neonates: A Review. Fetal and pediatric pathology, 39, 246–250.

Keidar, S., Gamliel-Lazarovich, A., Kaplan, M., Pavlotzky, E., Hamoud, S., Hayek, T., Karry, R. & Abassi, Z. (2005) Mineralocorticoid receptor blocker increases angiotensin-converting enzyme 2 activity in congestive heart failure patients. Circulation research, 97, 946–953.

Kim, C.J., Romero, R., Chaemsaithong, P., Chaiyasit, N., Yoon, B.H. & Kim, Y.M. (2015) Acute chorioamnionitis and funisitis: definition, pathologic features, and clinical significance. American journal of obstetrics and gynecology, 213, S29–52.

Knöfler, M., Haider, S., Saleh, L., Pollheimer, J., Gamage, T. & James, J. (2019) Human placenta and trophoblast development: key molecular mechanisms and model systems. Cellular and molecular life sciences : CMLS, 76, 3479–3496.

Kwon, J.Y., Romero, R. & Mor, G. (2014) New insights into the relationship between viral infection and pregnancy complications. American journal of reproductive immunology (New York, N.Y. : 1989), 71, 387–390.

Li, F. (2016) Structure, Function, and Evolution of Coronavirus Spike Proteins. Annual review of virology, 3, 237–261.

Li, G., He, X., Zhang, L., Ran, Q., Wang, J., Xiong, A., Wu, D., Chen, F., Sun, J. & Chang, C. (2020a) Assessing ACE2 expression patterns in lung tissues in the pathogenesis of COVID-19. J Autoimmun, 112, 102463–102463.

Li, M., Chen, L., Zhang, J., Xiong, C. & Li, X. (2020b) The SARS-CoV-2 receptor ACE2 expression of maternal-fetal interface and fetal organs by single-cell transcriptome study. PloS one, 15, e0230295.

Liu, X., Yin, S., Chen, Y., Wu, Y., Zheng, W., Dong, H., Bai, Y., Qin, Y., Li, J., Feng, S. & Zhao, P. (2018) LPS-induced proinflammatory cytokine expression in human airway epithelial cells and macrophages via NF-κB, STAT3 or AP-1 activation. Molecular medicine reports, 17, 5484–5491.

Livak, K.J. & Schmittgen, T.D. (2001) Analysis of relative gene expression data using real-time quantitative PCR and the 2(-Delta Delta C(T)) Method. Methods (San Diego, Calif.), 25, 402–408.

Lye, P., Bloise, E., Javam, M., Gibb, W., Lye, S.J. & Matthews, S.G. (2015) Impact of bacterial and viral challenge on multidrug resistance in first- and third-trimester human placenta. The American journal of pathology, 185, 1666–1675.

Lye, P., Bloise, E., Nadeem, L., Peng, C., Gibb, W., Ortiga-Carvalho, T.M., Lye, S.J. & Matthews, S.G. (2019) Breast Cancer Resistance Protein (BCRP/ABCG2) Inhibits Extra Villous Trophoblast Migration: The Impact of Bacterial and Viral Infection. Cells, 8.

Nadeem, L., Brkic, J., Chen, Y.F., Bui, T., Munir, S. & Peng, C. (2013) Cytoplasmic mislocalization of p27 and CDK2 mediates the anti-migratory and anti-proliferative effects of Nodal in human trophoblast cells. Journal of cell science, 126, 445–453.

Paquette, A.G., Shynlova, O., Kibschull, M., Price, N.D. & Lye, S.J. (2018) Comparative analysis of gene expression in maternal peripheral blood and monocytes during spontaneous preterm labor. American journal of obstetrics and gynecology, 218, 345.e341–345.e330.

Pringle, K., Tadros, M., Callister, R.J. & Lumbers, E. (2011a) The expression and localization of the human placental prorenin/renin-angiotensin system throughout pregnancy: Roles in trophoblast invasion and angiogenesis? Placenta, 32, 956–962.

Pringle, K.G., Tadros, M.A., Callister, R.J. & Lumbers, E.R. (2011b) The expression and localization of the human placental prorenin/renin-angiotensin system throughout pregnancy: roles in trophoblast invasion and angiogenesis? Placenta, 32, 956–962.

Qiao, J. (2020) What are the risks of COVID-19 infection in pregnant women? Lancet (London, England), 395, 760–762.

Rasmussen, S.A., Smulian, J.C., Lednicky, J.A., Wen, T.S. & Jamieson, D.J. (2020) Coronavirus Disease 2019 (COVID-19) and pregnancy: what obstetricians need to know. American journal of obstetrics and gynecology, 222, 415–426.

Redline, R.W., Faye-Petersen, O., Heller, D., Qureshi, F., Savell, V. & Vogler, C. (2003) Amniotic infection syndrome: nosology and reproducibility of placental reaction patterns. Pediatric and developmental pathology : the official journal of the Society for Pediatric Pathology and the Paediatric Pathology Society, 6, 435–448.

Romero, R., Salafia, C.M., Athanassiadis, A.P., Hanaoka, S., Mazor, M., Sepulveda, W. & Bracken, M.B. (1992) The relationship between acute inflammatory lesions of the preterm placenta and amniotic fluid microbiology. American journal of obstetrics and gynecology, 166, 1382–1388.

Salio, C., Lossi, L., Ferrini, F. & Merighi, A. (2005) Ultrastructural evidence for a pre- and postsynaptic localization of full-length trkB receptors in substantia gelatinosa (lamina II) of rat and mouse spinal cord. The European journal of neuroscience, 22, 1951–1966.

Schwartz, D.A. (2020) An Analysis of 38 Pregnant Women with COVID-19, Their Newborn Infants, and Maternal-Fetal Transmission of SARS-CoV-2: Maternal Coronavirus Infections and Pregnancy Outcomes. Archives of pathology & laboratory medicine.

Shah, P.S., Diambomba, Y., Acharya, G., Morris, S.K. & Bitnun, A. (2020) Classification system and case definition for SARS-CoV-2 infection in pregnant women, fetuses, and neonates. Acta obstetricia et gynecologica Scandinavica, 99, 565–568.

Shynlova, O., Nadeem, L., Zhang, J., Dunk, C. & Lye, S. (2020) Myometrial activation: Novel concepts underlying labor. Placenta, 92, 28–36.

Silasi, M., Cardenas, I., Kwon, J.Y., Racicot, K., Aldo, P. & Mor, G. (2015) Viral infections during pregnancy. American journal of reproductive immunology (New York, N.Y. : 1989), 73, 199–213.

Valdés, G., Neves, L.A., Anton, L., Corthorn, J., Chacón, C., Germain, A.M., Merrill, D.C., Ferrario, C.M., Sarao, R., Penninger, J. & Brosnihan, K.B. (2006) Distribution of angiotensin-(1-7) and ACE2 in human placentas of normal and pathological pregnancies. Placenta, 27, 200–207.

Vickers, C., Hales, P., Kaushik, V., Dick, L., Gavin, J., Tang, J., Godbout, K., Parsons, T., Baronas, E., Hsieh, F., Acton, S., Patane, M., Nichols, A. & Tummino, P. (2002) Hydrolysis of biological peptides by human angiotensin-converting enzyme-related carboxypeptidase. The Journal of biological chemistry, 277, 14838–14843.

Wan, Y., Shang, J., Graham, R., Baric, R.S. & Li, F. (2020) Receptor Recognition by the Novel Coronavirus from Wuhan: an Analysis Based on Decade-Long Structural Studies of SARS Coronavirus. Journal of Virology, 94, e00127–00120.

Wang, S., Guo, L., Chen, L., Liu, W., Cao, Y., Zhang, J. & Feng, L. (2020) A Case Report of Neonatal 2019 Coronavirus Disease in China. Clinical Infectious Diseases.

Zamaniyan, M., Ebadi, A., Aghajanpoor, S., Rahmani, Z., Haghshenas, M. & Azizi, S. (2020) Preterm delivery, maternal death, and vertical transmission in a pregnant woman with COVID-19 infection. Prenatal diagnosis.

Zeng, H., Xu, C., Fan, J., Tang, Y., Deng, Q., Zhang, W. & Long, X. (2020) Antibodies in Infants Born to Mothers With COVID-19 Pneumonia. Jama, 323, 1848–1849.

Zhang, H., Penninger, J.M., Li, Y., Zhong, N. & Slutsky, A.S. (2020a) Angiotensin-converting enzyme 2 (ACE2) as a SARS-CoV-2 receptor: molecular mechanisms and potential therapeutic target. Intensive Care Med, 46, 586–590.

Zhang, L., Jiang, Y., Wei, M., Cheng, B.H., Zhou, X.C., Li, J., Tian, J.H., Dong, L. & Hu, R.H. (2020b) [Analysis of the pregnancy outcomes in pregnant women with COVID-19 in Hubei Province]. Zhonghua fu chan ke za zhi, 55, 166–171.

